# Predictive model for the determination of caesarean sections in the Catalan National Health System: an approach based on clinical factors

**DOI:** 10.1101/2025.01.14.24319421

**Authors:** Albert Medina Català, Xavier Espada-Trespalacios, Maria Raventos Gil de Biedma, Assumpta Ricart Conesa, Roser Palau-Costafreda, Ramon Escuriet

## Abstract

**Objective:** To develop and validate a logistic regression model for analyzing the probability of caesarean section births, adjusted for clinical complexity, across public hospitals in Catalonia, and to identify deviations from expected caesarian section rates for benchmarking and quality improvement.

**Methods:** This retrospective cohort study analyzed data from the Catalan National Health System’s Minimum Basic Data Set (CMBD-AH), including all deliveries in public hospitals from January 2018 to June 2024. A logistic regression model was constructed using maternal and obstetric factors such as age, obstetric history, and clinical conditions. The model was validated through calibration plots and receiver operating characteristic (ROC) curve analysis, achieving an area under the curve (AUC) of 0.803.

**Results:** The analysis revealed variability in observed-to-expected caesarean section ratios across hospital complexity levels. Level III hospitals aligned closely with expected rates, reflecting adherence to clinical standards for high-complexity cases. Level I hospitals demonstrated significant variability, with 59.1% performing more cesareans than expected; smaller hospitals with fewer than 1,000 births exhibited the greatest deviation. The model highlighted both underperforming and overperforming institutions, offering actionable insights for resource allocation and policy interventions.

**Conclusions:** The logistic regression model provides a robust framework for evaluating caesarean section practices, enabling fair comparisons between hospitals by adjusting for clinical complexity. It supports the identification of non-clinical factors influencing cesarean practices and offers a critical tool for quality improvement and optimizing maternal healthcare within Catalonia’s public health system.

## BACKGROUND

Maternity and childbirth care are foundational elements of global health, significantly influencing maternal and neonatal outcomes. Across the globe, maternity care practices vary widely, reflecting differences in healthcare systems, socio-economic conditions, and cultural contexts. Over recent decades, a trend toward the medicalization of childbirth has emerged in industrialized countries (Filippini 2020; Van Teijlingen, 2004). This shift, motivated by the desire to mitigate maternal and neonatal mortality through medical technology, has led to increased interventions such as labour inductions and caesarean sections (Downe & Byrom, 2019; Renfrew et al., 2014). This medicalization of childbirth has been subject to criticism, particularly regarding the overuse of medical interventions and their interference with the physiological process of childbirth and the potential negative impact on the quality of care (Johanson et al., 2002; Miller et al., 2016).

One notable concern is the global rise in caesarean section rates (Betran et al., 2016; Angolile et al., 2023). Unnecessary caesarean sections are associated with increased risks for both mother and child, and long-term health impacts. Furthermore, the overuse of caesarean sections poses a significant challenge for healthcare system efficiency by driving up costs and straining resources. Addressing these concerns requires healthcare systems to strike a balance between clinical necessity and optimal health outcomes (Hoxha et al. 2021). In this context, healthcare financing models play an important role in promoting optimal clinical practice in a given context (Bailey et al., 2017). Payment systems that fail to consider patient complexity may unintentionally incentivize inappropriate caesarean section use, emphasizing the need for robust, complexity-adjusted indicators to guide equitable and effective care.

In Catalonia, a public health system characterized by universal access, maternity and postpartum care are provided in hospitals categorized into three levels based on their capacity to handle varying levels of maternal and neonatal complexity (Figure 1). Level I hospitals manage low-complexity cases, Level II hospitals handle intermediate cases, and Level III centres are equipped for high-complexity and high-risk cases. Despite this stratification, significant variability exists in caesarean section rates across hospitals, driven by differences in patient profiles, institutional practices, and resource availability. Observed caesarean rates often diverge from expected rates, highlighting the need for meticulous evaluation methods that incorporate clinical complexity.

**Figure 1.**
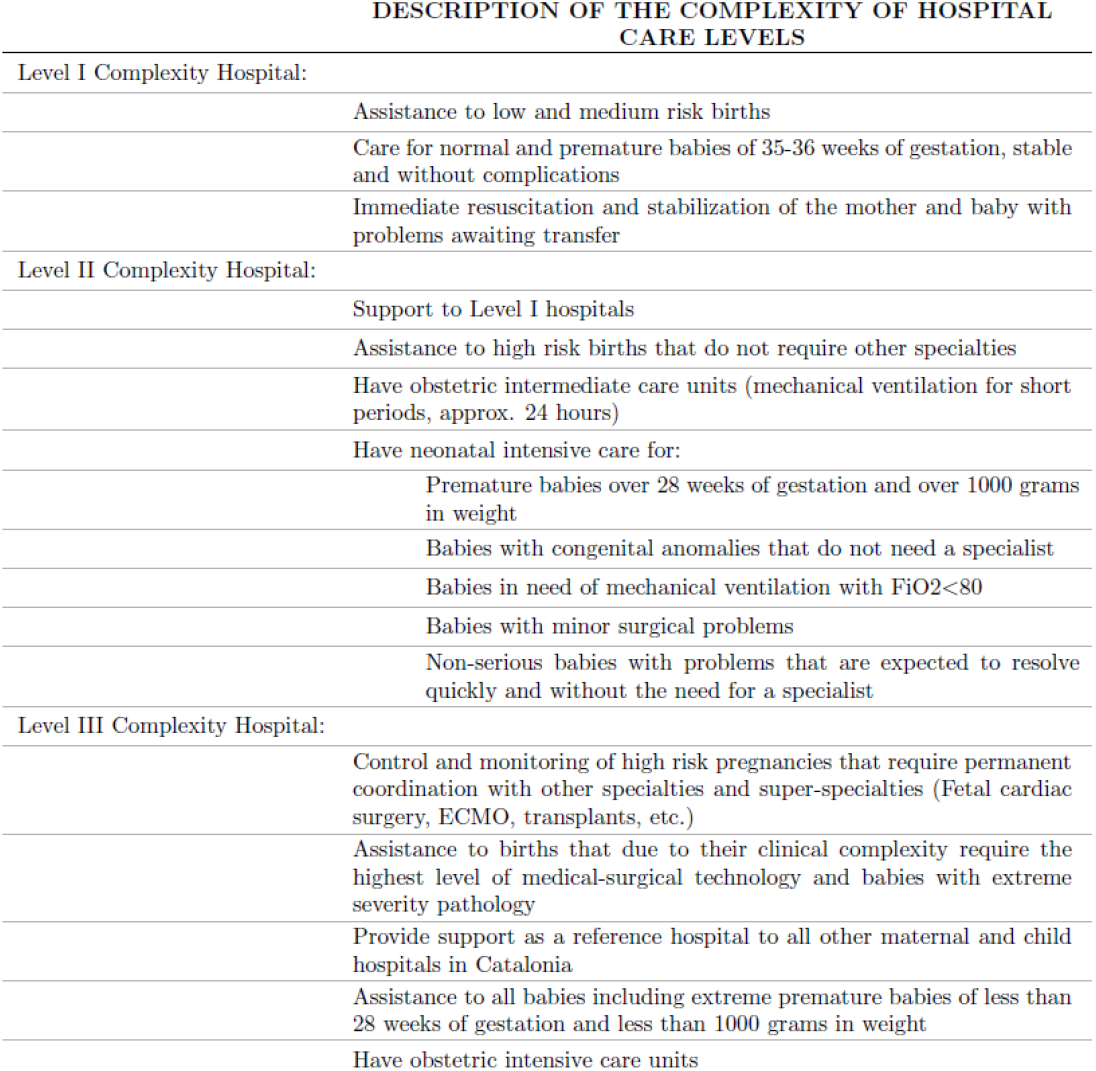
Description of the complexity of hospital care levels.

This study aims to address these gaps by developing a logistic regression model to analyse the probability of caesarean sections adjusted for clinical complexity. By focusing on observed and expected caesarean section rates across Catalonia, this research seeks to provide actionable insights to optimize healthcare practices, ensure equitable resource allocation, and align maternity care with clinical standards.

## METHODS

### Study Design

This study employed a retrospective cohort design to evaluate the probability of cesarean section births, adjusted for clinical complexity, across hospitals in Catalonia. The study aimed to develop and validate a logistic regression model capable of accounting for patient and hospital-specific characteristics, ensuring unbiased comparisons across institutions.

### Study Population

The study population included all women who gave birth in public hospitals within the Catalan Health System between January 2018 and June 2024. Women with incomplete records or stillbirths were excluded to maintain the integrity and consistency of the dataset.

### Data Collection

The data used in this study were sourced from the Catalan National Health System’s Minimum Basic Data Set (CMBD-AH). This dataset is maintained under strict regulatory and ethical standards and is fully anonymized before being made accessible. The data were analyzed internally within the Catalan Health Service to support health planning and service improvement. As the data were anonymized and did not involve identifiable information, the study did not require ethics committee approval or patient consent. This approach aligns with the standard procedures within the healthcare administration for systematically assessing and improving healthcare provider performance in Catalonia.

### Variables

The dependent variable in the model was the type of birth, categorized as either caesarean (1) or vaginal birth (0). Independent variables were selected based on their clinical relevance to caesarean likelihood and included maternal characteristics (age, obesity, and obstetric history); obstetric factors (Robson classification, prolonged labour, multiple gestation, foetal presentation abnormalities) and medical conditions (presence of preeclampsia/eclampsia, diabetes mellitus, low birth weight, placenta previa).

These variables were carefully defined and coded according to established clinical guidelines, ensuring consistency and reliability. To determine whether the births corresponded to nulliparous women, all births recorded since 2005 were analysed to ensure correct classification.

### Statistical Analysis

A binomial logistic regression model was used to estimate the probability of caesarean sections based on the independent variables. The model’s performance was evaluated using the area under the receiver operating characteristic (ROC) curve (AUC), which demonstrated an AUC of 0.803, indicating strong predictive capacity. All included variables were statistically significant, highlighting their relevance in predicting caesarean likelihood.

The model underwent internal validation to assess its reliability and accuracy. Calibration plots and ROC curves were used to evaluate goodness-of-fit and discriminative ability, confirming the model’s robustness across varying hospital levels and patient profiles.

The model facilitated the calculation of adjusted caesarean rates for individual hospitals, considering their specific case mix and complexity levels. Comparisons between observed and expected caesarean rates were conducted to identify deviations.

### Ethical Statement

The data were analyzed internally within the Catalan Health Service to enhance health planning. Given the non-identifiable nature of the data and its use for service improvement, ethics committee approval was not required. Confidentiality and adherence to ethical guidelines were strictly maintained throughout the study.

## RESULTS

The analysis of expected caesarean section rates across hospitals (Table 1), categorized by complexity levels, revealed significant trends and variability, with the hospital showing the lowest rate of expected caesareans at 17.4%, and the highest at 27.7%. High-Complexity Hospitals (Level III) exhibited the highest expected caesarean section rates, ranging from 23.8% at Hospital 41 to 27.7% at Hospital 38. These hospitals also handled the largest volume of births, with totals between 8,029 and 21,848. The consistently high expected rates align with their role in managing medically complex cases.

**Table 1.** Caesarean sections expected by hospitals and level of complexity.

| Hospital* | Complexity level** | Total births*** | % Expected CS |
| --- | --- | --- | --- |
| 38 | III | 18458 | 27,7% |
| 1 | III | 10469 | 27,3% |
| 39 | III | 21848 | 27,0% |
| 27 | III | 16281 | 26,3% |
| 24 | III | 8029 | 25,1% |
| 16 | III | 12463 | 24,9% |
| 15 | III | 8301 | 24,7% |
| 20 | II | 5405 | 24,2% |
| 8 | I | 7467 | 24,2% |
| 41 | III | 13208 | 23,8% |
| 36 | II | 14646 | 23,4% |
| 18 | II | 8363 | 23,2% |
| 17 | I | 7637 | 22,9% |
| 19 | II | 8294 | 22,9% |
| 4 | II | 5673 | 22,8% |
| 28 | I | 910 | 22,7% |
| 12 | II | 7196 | 22,4% |
| 2 | II | 5201 | 22,1% |
| 23 | I | 5854 | 22,1% |
| 14 | I | 780 | 22,0% |
| 40 | I | 3220 | 22,0% |
| 21 | I | 2534 | 21,3% |
| 9 | II | 6500 | 21,1% |
| 6 | II | 6110 | 21,1% |
| 11 | II | 6547 | 20,8% |
| 25 | II | 4884 | 20,8% |
| 37 | I | 2675 | 20,7% |
| 33 | I | 1865 | 20,6% |
| 7 | I | 4584 | 20,4% |
| 26 | I | 650 | 20,1% |
| 34 | I | 2737 | 20,0% |
| 29 | I | 3929 | 19,9% |
| 13 | I | 4504 | 19,8% |
| 31 | I | 3428 | 19,7% |
| 22 | I | 630 | 19,5% |
| 32 | I | 6833 | 18,6% |
| 3 | I | 2685 | 18,5% |
| 30 | I | 499 | 18,2% |
| 10 | I | 436 | 17,8% |
| 5 | I | 362 | 17,7% |
| 35 | I | 2632 | 17,4% |

\* Hospitals presented anonymized
\*\* Hospital level according to the complexity of the health care offered
\*\*\* Total births attended during the study period

| Hospital* | Complexity level** | Total births*** | % Expected CS |
| --- | --- | --- | --- |
| 38 | III | 18458 | 27,7% |
| 1 | III | 10469 | 27,3% |
| 39 | III | 21848 | 27,0% |
| 27 | III | 16281 | 26,3% |
| 24 | III | 8029 | 25,1% |
| 16 | III | 12463 | 24,9% |
| 15 | III | 8301 | 24,7% |
| 20 | II | 5405 | 24,2% |
| 8 | I | 7467 | 24,2% |
| 41 | III | 13208 | 23,8% |
| 36 | II | 14646 | 23,4% |
| 18 | II | 8363 | 23,2% |
| 17 | I | 7637 | 22,9% |
| 19 | II | 8294 | 22,9% |
| 4 | II | 5673 | 22,8% |
| 28 | I | 910 | 22,7% |
| 12 | II | 7196 | 22,4% |
| 2 | II | 5201 | 22,1% |
| 23 | I | 5854 | 22,1% |
| 14 | I | 780 | 22,0% |
| 40 | I | 3220 | 22,0% |
| 21 | I | 2534 | 21,3% |
| 9 | II | 6500 | 21,1% |
| 6 | II | 6110 | 21,1% |
| 11 | II | 6547 | 20,8% |
| 25 | II | 4884 | 20,8% |
| 37 | I | 2675 | 20,7% |
| 33 | I | 1865 | 20,6% |
| 7 | I | 4584 | 20,4% |
| 26 | I | 650 | 20,1% |
| 34 | I | 2737 | 20,0% |
| 29 | I | 3929 | 19,9% |
| 13 | I | 4504 | 19,8% |
| 31 | I | 3428 | 19,7% |
| 22 | I | 630 | 19,5% |
| 32 | I | 6833 | 18,6% |
| 3 | I | 2685 | 18,5% |
| 30 | I | 499 | 18,2% |
| 10 | I | 436 | 17,8% |
| 5 | I | 362 | 17,7% |
| 35 | I | 2632 | 17,4% |

\* Hospitals presented anonymized
\*\* Hospital level according to the complexity of the health care offered
\*\*\* Total births attended during the study period

Intermediate-Complexity Hospitals (Level II) reported expected caesarean section rates between 20.8% (Hospitals 25 and 11) and 24.2% (Hospital 20). Childbirth volumes in this group ranged from 4,884 to 14,646. While the expected rates were lower than those of Level III hospitals, they reflect the moderate complexity of cases managed by these institutions. Variability within this group suggests opportunities to refine clinical practices.

Low-Complexity Hospitals (Level I) displayed the greatest variability in expected caesarean section rates, which ranged from 17.4% at Hospital 35 to 24.2% at Hospital 8. Expected rates for some Level I hospitals approached those of Level II institutions, likely reflecting differences in patient profiles or local practices. Childbirth volumes in this group were notably smaller, spanning 362 to 7,637 births, with smaller hospitals exhibiting greater variability in expected rates.

Expected caesarean rates were generally found to correlate with hospital level, although exceptions were noted. For instance, Hospital 41, with an expected caesarean rate of 23.8%, falls significantly below the average for level III hospitals, which is 25.8%. Conversely, Hospital 8, classified as a Level I institution, reported an expected caesarean rate of 24.2%, notably above the average of 20.3% for hospitals at this level. However, Hospital 35, also Level I, reported the lowest expected rate at 17.4%. Similarly, Hospital 20, with an expected rate of 24.2%, represents the level II hospital with the highest rate, exceeding the level II average of 22.3%.

Figure 2 illustrates the relationship between observed and expected caesarean rates based on the clinical complexity of each hospital, grouped by care level. Values above 1.0 indicate that a hospital performed more caesareans than expected for its clinical complexity. Conversely, values below 1.0 suggest fewer caesareans than expected. The findings reveal significant variability across hospitals, with a general trend towards a higher proportion of caesareans in hospitals with lower care levels.

**Figure 2.**
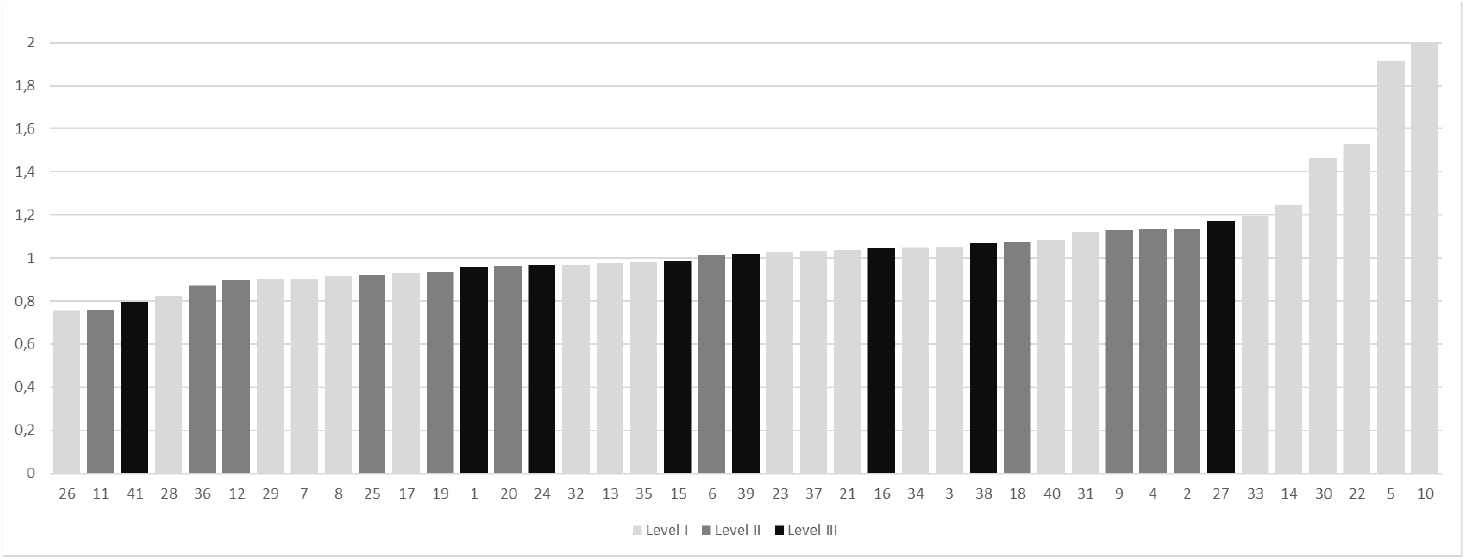
Observed and expected caesarean sections according to the model by hospital and level of complexity

Level III Hospitals demonstrated the closest alignment with expected caesarean rates. Observed-to-expected ratios for these hospitals ranged from 0.79 to 1.17, with an average ratio of 1.0, indicating that, overall, these hospitals performed caesareans as expected given the complexity of the cases they manage. This group demonstrated the closest alignment with the expected norm. Level II Hospitals, exhibited slightly more variability in their observed-to-expected ratios, which ranged from 0.76 to three hospitals reaching a maximum ratio of 1.13. The average ratio for this group was 0.98, with a standard deviation of 0.12, with no significant association between the number of births and deviations from the expected observed-to-expected caesarean ratio in level II hospitals.

In the analysis of level I hospitals, 59.1% were found to perform more caesareans than expected for their level of clinical complexity. Of these, 71.4% were hospitals with fewer than 1,000 births during the study period. It is also notable that this group exhibited the highest standard deviation (0.32) among the three levels, indicating greater variability in the observed-to-expected caesarean ratio for hospitals with this care level.

## DISCUSSION

The findings of this study provide significant insights into cesarean section practices across hospitals in Catalonia, highlighting the importance of clinical complexity in explaining variability in caesarean section rates (Bragg et al., 2010). The observed-to-expected ratios demonstrated alignment with expected norms for Level III hospitals, suggesting that high-complexity institutions generally adhere to appropriate clinical guidelines. This consistency reflects the alignment of these hospitals’ practices with their role as referral centers for high-risk cases. Conversely, the variability observed in Level I hospitals raises concerns about potential overuse of cesareans, with ratios exceeding expected values. Such trends emphasize the influence of non-clinical factors, including resource availability, staffing, models of care, or institutional policies (Palau-Costafreda et al., fc). These findings highlight the need for targeted interventions to ensure practices align with clinical standards while avoiding unnecessary medical interventions.

These findings align with existing literature on the impact of hospital characteristics and payment systems on cesarean section rates (Bailey et al. 2017). As a significant global public health concern, understanding the relationship between payment systems and the rate of cesarean deliveries is crucial for policymakers aiming to optimize maternal and neonatal outcomes. Research has consistently shown that the mode of hospital reimbursement influences the frequency of medical procedures, including cesarean births (Bailey et al., 2017; Main et al., 2019). Fee-for-service models, for instance, have been associated with higher CS rates, potentially due to financial incentives for performing more procedures. On the other hand, capitated models, while effective at containing costs, may inadvertently discourage medically necessary CSs. Payment systems emphasizing quality outcomes offer a balanced approach by aligning financial sustainability with evidence-based clinical practices.

In this context, there is a pressing need to develop hospital payment policies that incentivize appropriate use of CSs without compromising patient autonomy or clinical judgment (Zhou et al.). Proposed interventions include the implementation of bundled payments with integrated quality measures to discourage unnecessary cesareans, a greater emphasis on pay-for-performance systems that reward adherence to clinical guidelines, and increased transparency in hospital billing practices to empower patient choice.

To address these systemic issues, several policy recommendations emerge from the findings. Bundled payment systems with quality measures could discourage unnecessary cesareans, while pay-for-performance models promoting adherence to clinical guidelines may encourage optimal care.

Enhanced transparency in hospital billing practices could empower patients and foster greater accountability within the healthcare system. By incorporating these strategies, policymakers can optimize maternal and neonatal health outcomes while ensuring financial sustainability.

A previous study in Catalonia has already demonstrated that the volume of deliveries attended in a hospital and the type of financing significantly influence the incidence of cesarean births (Escuriet et al., 2014). This study confirms these findings in the Catalan context while contributing a novel tool in the form of a logistic model for analyzing cesarean practices. This complexity-adjusted model facilitates fairer comparisons between hospitals and has the potential to support hospital payment models without negatively impacting clinical practice. By accounting for differences in patient case mix, this model adds value to Catalonia’s public health toolkit.

As demonstrated, Level III hospitals performed as expected, validating the model’s applicability in high-complexity settings. Meanwhile, the tool highlights deviations in Level I and II hospitals, offering actionable insights for targeting areas of improvement. These findings emphasize the importance of integrating clinical complexity into evaluations of cesarean practices to improve healthcare quality and equity. By aligning clinical practices with objective standards, the healthcare system can optimize maternal and neonatal outcomes while ensuring sustainable and equitable care delivery.

### Strengths and limitations

This study leveraged a large and reliable dataset from the Catalan Health System (CMBD-AH), enabling robust analysis and adjustment for clinical complexity, which allowed unbiased comparisons between hospitals. The logistic regression model demonstrated strong predictive power with an AUC of 0.803, providing actionable insights for benchmarking healthcare practices. However, the retrospective design relied on historical data, potentially missing undocumented factors influencing caesarean decisions, such as patient preferences or institutional policies. The absence of external validation limits generalizability beyond Catalonia, and the exclusion of private hospitals reduces applicability across all healthcare settings. Additionally, unmeasured contextual factors, such as staffing levels or cultural attitudes, could also affect caesarean rates but were not included in the model. Despite these limitations, the study provides a thorough approach to evaluating caesarean section practices within the public healthcare system.

### Implications for practic

The findings of this study emphasize the urgent need for policy interventions that incentivize the appropriate use of cesarean sections without compromising patient autonomy or clinical judgment. Implementing bundled payment systems with integrated quality measures could play a role in discouraging unnecessary cesareans while ensuring financial sustainability and adherence to clinical guidelines. Additionally, these payment models could enhance transparency in hospital practices, empowering patients and fostering accountability within the healthcare system. By aligning financial incentives with evidence-based practices, policymakers can drive improvements in maternal and neonatal outcomes while optimizing resource use across Catalonia’s public healthcare system.

## Conclusions

The logistic regression model developed in this study serves as a powerful statistical tool for quantifying the probability of cesarean delivery, adjusting for the clinical complexity of each case. By incorporating clinical factors such as maternal age, obstetric history, and relevant medical variables, the model provides an accurate approximation of how these factors influence medical decision-making. This nuanced analysis allows the differentiation between medically necessary cesareans and those that may not be justified on clinical grounds.

The results of this model provide a robust instrument for benchmarking within Catalonia’s integrated health system (SISCAT). By adjusting cesarean section rates for clinical complexity, the model facilitates the identification of hospitals with practices that deviate from expected standards. For hospitals with significantly higher or lower cesarean rates after adjustment, further analysis can determine whether these deviations are driven by clinical needs or other non-clinical factors, such as organizational pressures, patient preferences, or systemic inefficiencies. This information is pivotal for improving the quality of care, aligning obstetric practices with objective medical criteria, and optimizing cesarean use within the public health system.

Moreover, the logistic regression model offers a reliable framework for optimizing resource allocation across Catalonia’s public healthcare system. By identifying deviations in cesarean rates, this study provides actionable insights that enable the alignment of maternal healthcare practices with objective clinical standards, ensuring equitable and effective care delivery. Ultimately, the model fosters a deeper understanding of decision-making in obstetrics, supporting both quality improvement initiatives and evidence-based policymaking.

## Data Availability

All data produced in the present study are available upon reasonable request to the authors

## FUNDING

No funding was received for this study.

## DECLARATION OF COMPETING INTEREST

The authors declare that they have no known competing financial interests or personal relationships that could have appeared to influence the work reported in this paper.

## Notes

### Competing Interest Statement

The authors have declared no competing interest.

### Funding Statement

This study did not receive any funding

### Author Declarations

The study used only openly available human data sourced from the Catalan National Health System's Minimum Basic Data Set (CMBD-AH). This dataset is maintained under strict regulatory and ethical standards and is fully anonymized before being made accessible. The data were analyzed internally within the Catalan Health Service to support health planning and service improvement. As the data were anonymized and did not involve identifiable information, the study did not require ethics committee approval or patient consent. This approach aligns with the standard procedures within the healthcare administration for systematically assessing and improving healthcare provider performance in Catalonia.

